# Mobility data can explain the entire COVID-19 outbreak course in Japan

**DOI:** 10.1101/2020.04.26.20081315

**Authors:** Junko Kurita, Tamie Sugawara, Yasushi Ohkusa

## Abstract

**Background:** In Japan, as a countermeasure against the COVID-19 outbreak, voluntary restrictions against going out (VRG) have been applied.

**Object:** We examined mobility information provided by Apple Inc. to a susceptible–infected–recovery model.

**Method:** When applying a polynomial function to daily Apple data with the SIR model, we presumed the function up to a cubic term as in our earlier study.

**Results:** Estimation results demonstrated R_0_ as 1.507 and its 95% confidence interval was [1.502, 1.509].. The estimated coefficients of Apple data was 1.748 and its 95% confidence interval was [1.731, 1.788].

**Discussion and Conclusion:** Results show that mobility data from Apple Inc. can explain the entire course of the outbreak in COVID-19 in Japan. Therefore, monitoring Apple data might be sufficient to adjust control measures to maintain an effective reproduction number of less than one.

## Introduction

Some reasons for how the COVID-19 outbreak peak can be overcome might be inferred to support planning and evaluation. Two modes are herd immunity [1] and infection countermeasures [2].

In Japan, in preference to lockdowns such as those instituted in European and North American countries, voluntary restrictions against going out (VRG) were announced by national and local governments from the end of March [3]. However, the VRG program intensity has been changing over time. Public cooperation with VRG has also been changing. Lockdowns such as those instituted in Europe and North America might eventually lead to cessation as determined by their respective governments. However, in Japan, efforts are voluntary: people can adjust their own degree of cooperation independently. Therefore, the government should monitor current circumstances of an outbreak when moderating requirements for VRG. However, some delay occurred in reporting the number of the newly infected patients. One delay is attributable to the incubation period from infection to onset; another delay is that occurring after onset until reporting. Taken together, for approximately two weeks, one is unable to observe a precise daily number of newly infected persons. Therefore, because the latest information might not be timely, policies might be less effective if a government waits two weeks to conduct decision-making.

To date, susceptible–infected–recovery (SIR) models for COVID-19 incorporating countermeasures have emphasized the date of countermeasure initiation [2,4,5]. However, the use of VRG has expanded gradually, at least in the case of Japan. Therefore, an all-or-nothing approach such as that implied by an SIR model might be inappropriate. A more continuous variable is expected to be necessary to assess compliance with VRG over time.

Those variables have been reported from several services, including those of Apple Inc. and Alphabet Inc. (hereinafter Apple and Google, respectively) worldwide, and Nippon Telegraph and Telephone (NTT) and East and West Japan Railway companies (JR) in Japan. Of those, Apple, as the front-runner of this service, has provided related data from January 13. The daily information has included the number of trips from home by type, such as driving and walking [6].

Using mobility information provided by Apple, we examined associations with the entire course of the outbreak in Japan. It might be sufficient to monitor the mobility data to control it if one can explain the entire course of the outbreak solely on the basis of mobility data.

## Methods

We applied a simple deterministic SIR model [2,4,5] along with mobility data to the epidemic curve in Japan, with its 120 million population. We assume an incubation period that conforms to the empirical distribution in Japan. The number of symptomatic patients reported by the Ministry of Labour, Health and Welfare (MLHW) for January 14 – May 24 published [7] on May 25 were used. Some patients were excluded from data: those presumed to be persons infected abroad or infected as passengers on the Diamond Princess. Those patients were presumed not to represent community-acquired infection in Japan.

For onset dates of some symptomatic patients that were unknown, we estimated their onset date from an empirical distribution with duration extending from onset to the report date among patients for whom the onset date had been reported. We estimated the onset date of patients for whom onset dates were not reported as follows: Letting *f*(*k*) represent this empirical distribution and letting *N_t_* denote the number of patients for whom onset dates were not published by date *t*, then the number of patients for whom the onset date was known is *t*-1. The number of patients for whom onset dates were not available was estimated as *f*(1)*N_t_*. Similarly, the number of patients with onset date *t-*2 and for whom onset dates were not available was estimated as *f*(2)*N_t_*. Therefore, the total number of patients for whom the onset date was not available, given an onset date of *s*, was estimated as 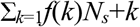 for the long duration extending from *s*.

Moreover, the reporting delay for published data from MHLW might be considerable. In other words, if *s+k* is larger than that in the current period *t*, then *s+k* represents the future for period *t*. For that reason, *Ns+k* is not observable. Such a reporting delay engenders underestimation bias of the number of patients. For that reason, it must be adjusted as 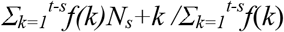. Similarly, patients for whom the onset dates were available are expected to be affected by the reporting delay. Therefore, we have 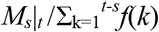, where *M_s_*|*t* represents the reported number of patients for whom onset dates were within period *s*, extending until the current period *t*.

We defined R(*t*) as the number of infected patients on day *t* divided by the number of patients who were presumed to be infectious. The number of infected patients was calculated from the epidemic curve by the onset date using a distribution of the incubation period. The distribution of infectiousness in symptomatic and asymptomatic cases was assumed to be 30% on the onset day, 20% on the following day, and 10% for the subsequent five days [8].

To clarify associations among the epidemic curve and Apple data, we incorporated a polynomial function of daily Apple data into the SIR model. Especially, we presumed a function up to cubic term as in our earlier study [9]. For analyses of the present study, R_0_ was defined as the average secondary infection from one infected person if all persons are susceptible and if their degree of mobility level is normal, i.e. equal to that of January 13, 2020.

We sought R_0_ and the cubic function of Apple data to fit the epidemic curve by minimizing the sum of the absolute difference. Its 95% confidence interval (CI) was calculated using 10,000 iterations of bootstrapping for the empirical distribution of epidemic curves.

### Ethical consideration

All information used for this study has been published [6,7]. There is therefore no ethical issue related to this study.

## Results

As of May 25, using data for January 14 – May 25 in Japan, 14,972 community-acquired cases were identified, excluding asymptomatic cases. Figure 1 presents an empirical distribution of the duration of onset to reporting in Japan. The maximum delay was 30 days. Figure 2 depicts the empirical distribution of incubation periods among 125 cases for which the exposed date and onset date were published by MHLW in Japan. The mode was six days. The average was 6.6 days.

**Figure 1:**
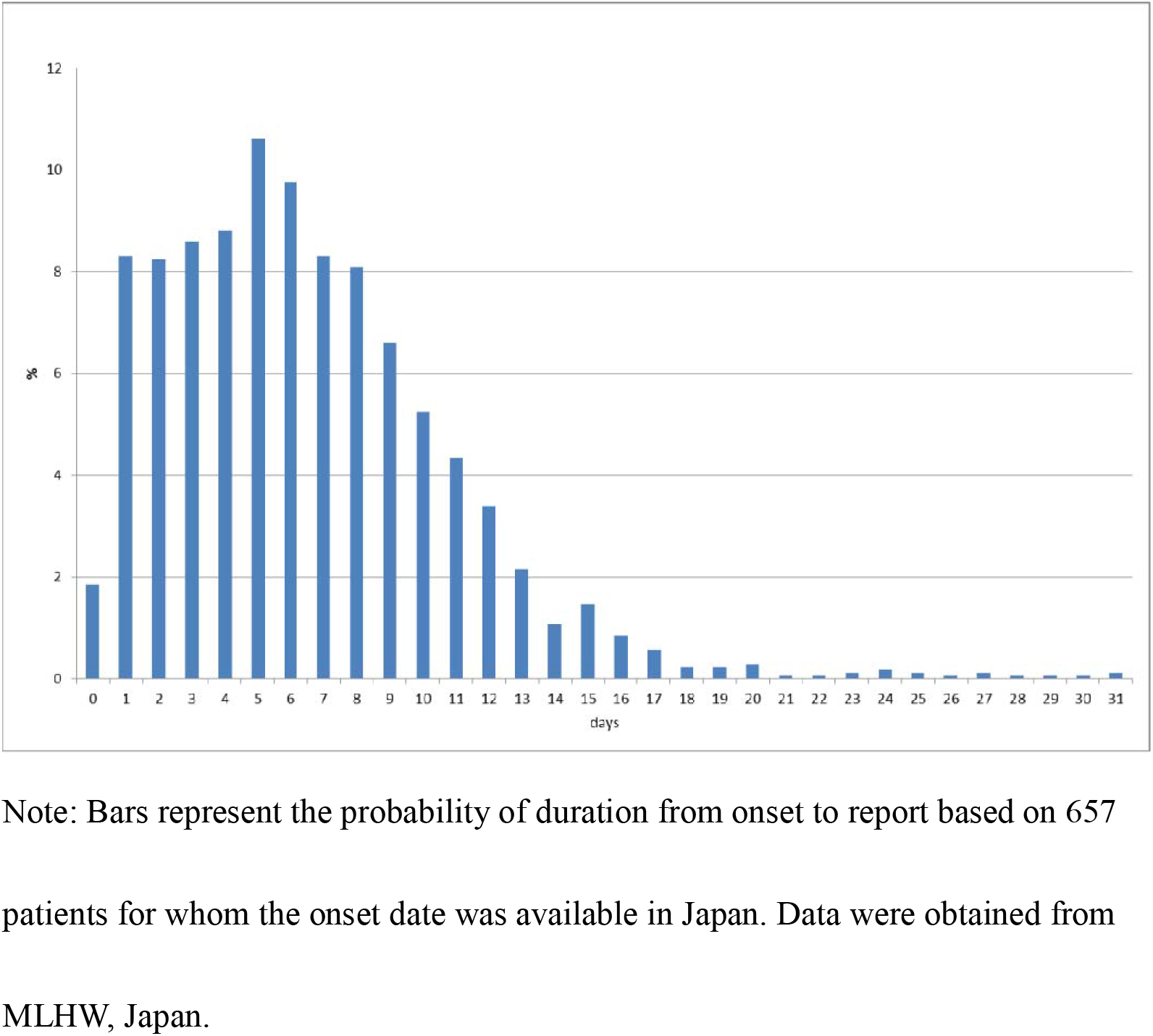
Empirical distribution of duration from onset to report by MLHW, Japan.

Figure 3 depicts the epidemic curves, the fitted line, and its 95% confidential interval. Estimation results showsthat R_0_ was 1.507 and its 95% confidence interval (CI) was [1.502, 1.509]. The estimated coefficients of Apple data was 1.748 and its 95% CI was [1.731, 1.788]. The range of the 95% CI was a small area that covered almost the entire epidemic curve, as shown in Figure 3.

**Figure 2:**
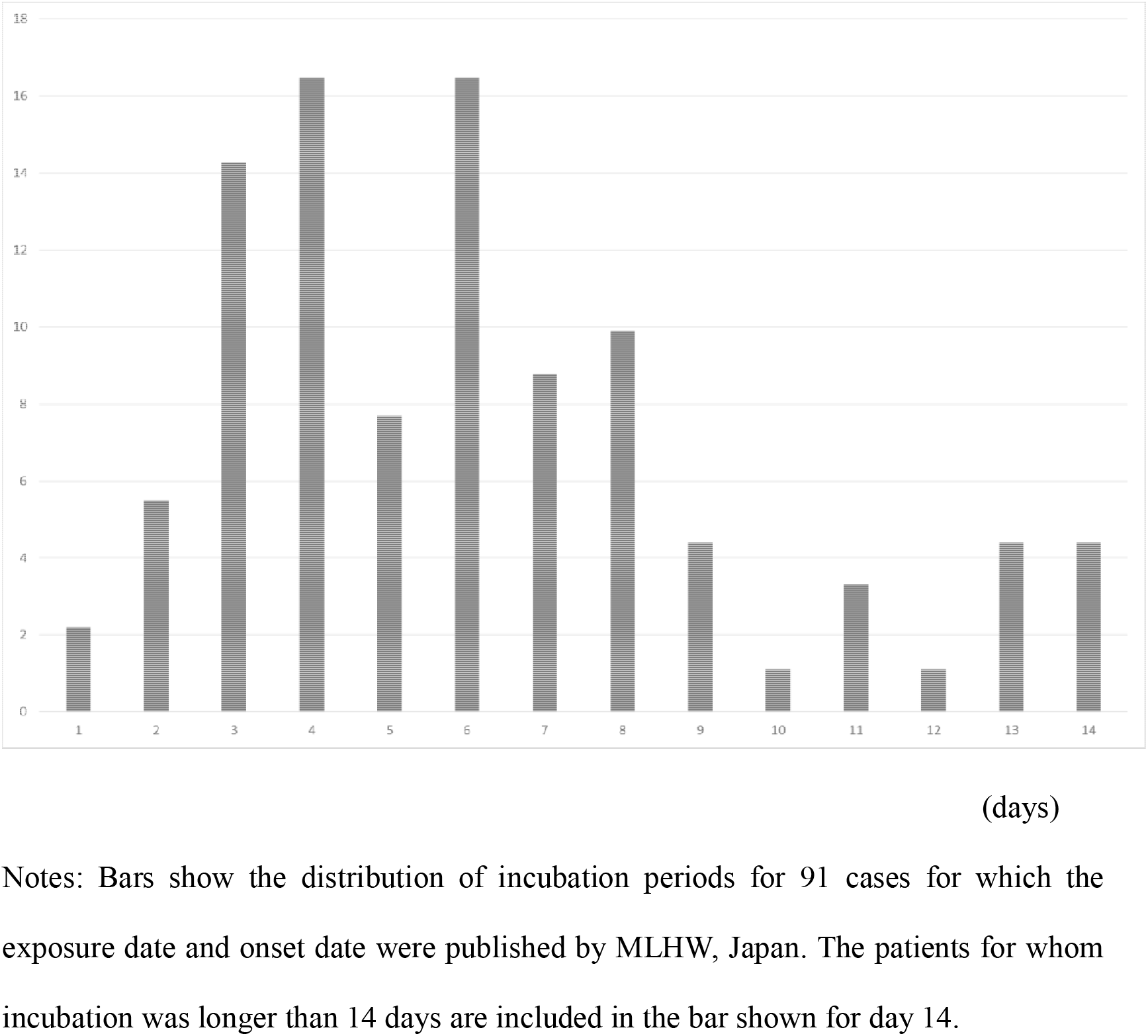
Empirical distribution of the incubation period published by MLHW, Japan.

**Figure 3:**
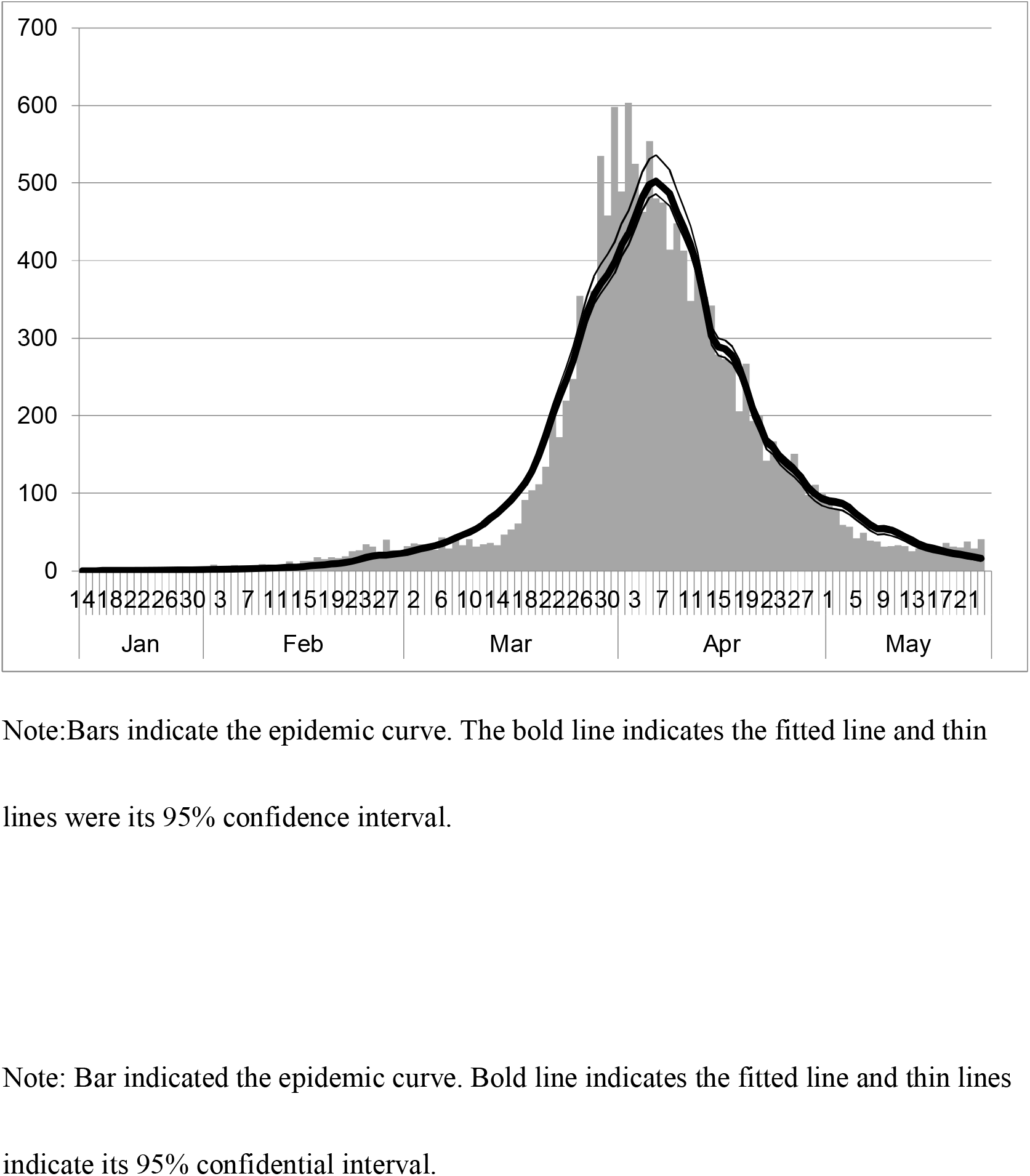
Epidemic curve, the fitted line and its confidence interval

## Discussion

One can reasonably infer that the four parameters explained the entire course of the outbreak completely. In other words, the variation of mobility data can include data for the outbreak. Therefore, the monitoring of Apple data and countermeasures depend on their probable capability of controlling the outbreak, at least in Japan. Actually, Apple data were provided with a two-day delay. However, we can infer the current situation of the newly infected in just two days. The average incubation period and reporting delay were 6–7 days, respectively. Therefore the delay from new infection to reporting was almost two weeks. Results suggest that Apple data are the most timely and precise information sources for newly infected persons.

In this study, R_0_ was smaller than that reported from an earlier study [10–12]: the estimated R_0_ for COVID-19 is 2.24–3.58. However, one study revealed R_0_ in Japan through 26 February as just 0.6 [13], meaning that an outbreak would never occur in Japan. Such estimates might also mislead policies for countermeasures in Japan, which have so far necessitated adherence to contact tracing to detect clusters. In both cases, one must be reminded that R_0_ in this study was defined as normal mobility equal to that of January 13, 2020. Therefore, it might be difficult or meaningless to compare R_0_ with similar figures referred from other reports.

The present study has some limitations. First, we examined the explanatory power for the COVID-19 outbreak nationwide, but is it applicable at the prefecture level? For example, did Apple data for Osaka prefecture have sufficient explanatory power to assess the outbreak dynamics? If so, one must monitor Apple data from Osaka or other prefectures carefully. Similarly, are these data and methods useful for countries other than Japan? If these methods hold in the US, for example, Apple data might contribute to efforts at controlling outbreaks there.

Secondly, one must be reminded that Apple data show a proportion of users leaving their residence. The data do not directly indicate a number of contacts or even a rate of contact. In other words, Apple data reflect no intensity of the respective contacts. However, such measurement of contact intensity is extremely difficult. That measurement represents a future research objective.

Thirdly, although Apple data was better than others, the users of Apple products and services might be limited to young or healthy persons. By contrast, information from NTT or JR might reflect the activities of an otherwise limited scope or number of users. Therefore, a combination of these data with Apple data might be better than the data used for the present study. That is anticipated as our next challenge.

## Conclusion

We demonstrated that mobility data from Apple can explain the entire course of the outbreak in COVID-19 in Japan. Therefore, it might be sufficient to monitor Apple data to adjust control measures to maintain the effective reproduction number as less than one.

## Data Availability

1.Japan Ministry of Health, Labour and Welfare. Press Releases.
2.Apple. Mobility trend Data.

https://www.mhlw.go.jp/stf/newpage_10723.html

https://www.apple.com/covid19/mobility

## Acknowledgments

We acknowledge the great efforts of all staff at public health centers, medical institutions, and other facilities who are fighting the spread and destruction associated with COVID-19.

